# Preoperative atelectasis in patients with obesity undergoing bariatric surgery: a cross-sectional study

**DOI:** 10.1101/2024.01.11.24301138

**Authors:** Javier Mancilla-Galindo, Jesus Elias Ortiz-Gomez, Orlando Rubén Pérez-Nieto, Audrey De Jong, Diego Escarramán-Martínez, Ashuin Kammar-García, Luis Carlos Ramírez Mata, Adriana Mendez Díaz, Manuel Alberto Guerrero-Gutiérrez

## Abstract

**Background:** Pulmonary atelectasis is present even before surgery in patients with obesity. No study has reported the prevalence of preoperative atelectasis in obese patients to date. We aimed to estimate the prevalence and extension of preoperative atelectasis in patients with obesity undergoing bariatric surgery and to determine if variation in preoperative SpO2 values in the seated position at room air is explained by the extent of atelectasis coverage in the supine position.

**Methods:** Cross-sectional study in a single center specialized in laparoscopic bariatric surgery. Preoperative chest computed tomographies were reassessed by a senior radiologist to quantify the extent of atelectasis coverage as a percentage of total lung volume. Patients were classified as having atelectasis when the affection was ≥2.5%, to estimate the prevalence of atelectasis. Crude and adjusted prevalence ratios (PR) and odds ratios (OR) were obtained to assess the relative prevalence of atelectasis and percentage coverage, respectively, with increasing obesity category. Inverse probability weighting was used to assess the total, direct (not mediated), and indirect (mediated through atelectasis) effects of BMI on preoperative SpO2, and to quantify the magnitude of mediation (proportion mediated).

**Results:** In 236 patients with a median BMI of 40.3 kg/m^2^ (IQR: 34.6–46.0, range: 30.0–77.3), the overall prevalence of atelectasis was 32.6% (95%CI: 27.0–38.9) and by BMI category: 30-35 kg/m^2^, 12.7% (95%CI: 6.1–24.4); 35-40 kg/m^2^, 28.3% (95%CI: 17.2–42.6); 40-45 kg/m^2^, 12.3% (95%CI: 5.5–24.3); 45-50 kg/m^2^, 48.4% (95%CI: 30.6–66.6); and ≥50 units, 100% (95%CI: 86.7–100). Compared to the 30-35 kg/m^2^ group, only the categories with BMI ≥45 kg/m^2^ had significantly higher relative prevalence of atelectasis — 45-50 kg/m^2^, aPR=3.52 (95%CI: 1.63–7.61) and ≥50 kg/m^2^, aPR=8.0 (95%CI: 4.22–15.2) — and higher odds of greater atelectasis percentage coverage: 45–50 kg/m^2^, aOR=7.5 (95%CI: 2.7–20.9) and ≥50 kg/m^2^, aOR=91.5 (95%CI: 30.0–279.3). Atelectasis percent alone explained 70.2% of the variation in preoperative SpO2. The proportion of the effect of BMI on preoperative SpO2 values <96% mediated through atelectasis was 81.5% (95%CI: 56.0–100).

**Conclusions:** The prevalence and extension of atelectasis increased with higher BMI, being significantly higher at BMI ≥45 kg/m^2^. Preoperative atelectasis mediated the effect of BMI on SpO2 at room air in the seated position. The high prevalence of atelectasis before surgery and their impact on SpO2 could be important factors to consider when deciding ventilation strategies during surgery and for the interpretation of the clinical significance of postoperative atelectasis.

**Key points:**

- Question: What is the prevalence of preoperative atelectasis in patients undergoing bariatric surgery and are changes in the preoperative peripheral saturation of oxygen (SpO2) at room air in the seated position explained by the extent of atelectasis coverage on chest CT in the supine position?
- Findings: Preoperative atelectasis were highly prevalent (32.6%, 95%CI: 27.0–38.9) in patients with obesity and a BMI above 45 kg/m^2^ was associated with a higher relative prevalence and atelectasis percentage coverage, the latter of which alone explained 70.2% of the variation in SpO2, with 81.5% (95%CI: 56.0-100) of the effect of BMI on SpO2 <96% mediated through atelectasis.
- Meaning: Pulmonary atelectasis are detectable before surgery in obese patients and largely explain decreased preoperative SpO2 values, which could be an important factor to consider when interpreting postoperative atelectasis and for deciding perioperative ventilation strategies.

## INTRODUCTION

North-American countries have a high prevalence of obesity in adults: USA,^1^ 41.9%; Canada,^2^ 30.0%; and Mexico,^3^ 36.9%. People living with obesity are more susceptible to complications during the perioperative period due to factors such as reduced functional residual capacity, cephalic displacement of the diaphragm, and increased adipose tissue in the chest wall and abdomen.^4,5^ Increasing body mass index (BMI) is associated with a greater decline in lung vital capacity during anesthesia.^6^

Obesity is an important risk factor for lung complications (i.e., atelectasis) in patients undergoing anesthesia, as these patients suffer from mechanical compression (leading to airway narrowing and closure), obstructive sleep apnea (OSA), and obesity hypoventilation syndrome.^7^ Perioperative atelectasis is more common in patients with obesity compared to patients with a normal BMI, with the former experiencing persistence of atelectasis 24 hours after surgery.^8^ Although attention has focused on postoperative atelectasis, Lagier et al. recently highlighted that “the direct impact of intraoperative pulmonary atelectasis on postoperative outcomes is still unclear”.^9^

Besides the previously mentioned mechanisms, altered lung surfactant production induced by obesity could cause atelectasis, as animal models with obesity have demonstrated surfactant deficiency relative to alveolar surface area.^10,11^ Obese patients with asthma have reduced surfactant protein (SP)-A levels,^12^ a mechanism that could increase surface tension, thereby facilitating alveolar collapse.^13^ Notably, weight loss after bariatric surgery has been shown to improve lung function due to the normalization of SP-A and SP-C expression.^14^

Despite being less studied, atelectasis occur even before surgery in patients with obesity.^8,15^ Our hypotheses were that the occurrence of atelectasis increases with a higher degree of obesity and that the biological effect of BMI on preoperative peripheral saturation of oxygen (SpO2) is mediated by atelectasis, despite these measurements being taken in different positions (seated vs supine). Thus, the objective of this study was to assess the prevalence and extension of preoperative atelectasis (primary outcome) in patients with increasing degrees of obesity (exposure) and to assess the extent to which the effect of BMI on SpO2 (secondary outcome) is mediated through atelectasis.

## METHODS

### Study Design and Setting

This was a single-center cross-sectional study conducted in a specialized center for laparoscopic bariatric surgery in Tijuana, Mexico mainly receiving patients from abroad. The study period was the month of June 2020. Adult patients who presented for elective bariatric surgery and underwent chest CT scan screening for coronavirus disease (COVID-19) were eligible. Exclusion criteria were a COVID-19 Reporting and Data System (CO-RADS)^16^ score of 3 to 6, positive antibody test against SARS-CoV-2, prior history of COVID-19, neuromuscular disease, or bronchiectasis. This study was reviewed and approved by the ethics committee of Hospital General de Tijuana (CONBIOÉTICA-02-CEI-001-20170526, approval no. 001771) and the requirement for written informed consent was waived by the IRB. Greater details on the COVID-19 screening algorithm, study setting, and sample size estimation are provided as **Supplemental Methods**.

### Exposure

Weight was measured in a 90x90 cm platform scale (Rhino^©^ PLABA-9) with a maximal capacity of 1,000 kg and precision of 200 grams. For height determination, patients are encouraged to stand with the heels together and buttocks, shoulders, and head in contact with a stadiometer (precision 0.1 mm). BMI (kg/m^2^) was determined as the ratio of weight (kilograms) and squared height (m^2^). Obesity class categories were created from BMI according to WHO criteria:^17^ class 1 (30–35 kg/m^2^), 2 (35–40 kg/m^2^), and 3 (≥40 kg/m^2^). Class 3 obesity subgroups were further defined as: 40–45 kg/m^2^, 45–50 kg/m^2^, and ≥50 units.

### Outcomes

The main outcome was the prevalence of atelectasis. Secondary outcomes were the degree of atelectasis coverage as a percentage of lung volume and SpO2 during the preanesthetic assessment.

High-resolution chest CT images (1mm slices, 120kV, 50mA, scan time: 0.5sec, FOV L: 240) were obtained with a Toshiba^©^ Aquilion 16 Slice CT Scanner and archived in EvoView PACS (U.M.G. Inc.). A senior radiologist was blinded to the patient’s BMI and assessed the presence and extent of atelectasis (OsiriX^©^ viewer) by measuring the total area of the lung—pixels with density values between –1000 and +100 Hounsfield Units (HU)—. Densities considered to indicate atelectasis were identified in dependent lung regions and calculated by including all pixels —HU between –100 and +100—.^18^ Because a median atelectasis percentage coverage of 2.5% has been shown to have low-to-no impact on oxygenation,^19^ percentage of atelectasis coverage was registered by rounding to the lower 2.5% category (i.e. values <2.5% were rounded to 0%). Thus, all patients with an atelectasis percentage ≥2.5% were classified as having atelectasis. We further recorded the predominance of atelectasis on chest CT as having right lung base predominance or affecting bilateral lung bases.

SpO2 was determined during the preanesthetic assessment with the patient seating, at rest, and at room air (FiO2: 21%) with a pulse oximeter (Masimo SET®) with precision of 2% in the range of 70-100%.

### Confounders

All hypothesized relationships between the exposure, mediators, outcomes, and confounders were defined a priori and schematized in a directed acyclic graph (DAG) with the *DAGitty* software^20^ according to published literature (**Supplemental Methods**). The diagram was updated by testing implied conditional independencies as described by Ankan, et al.^21^ (**Supplemental Figure 1**).

Age, sex, and mean altitude of the state of residence in meters above sea level (m.a.s.l.) constituted the minimal set of adjustment for the relationship between obesity class and atelectasis. For the relationship between BMI and preoperative SpO2, atelectasis percentage was studied as the mediator of the effect. The minimal set of confounders for BMI→SpO2 was the same as above, whereas the set of confounders for atelectasis→SpO2 were age, sex, BMI, altitude, asthma, OSA, and chronic obstructive pulmonary disease (COPD) (**Supplemental Figures 2A and 2B**). These were used for the obtention of inverse probability weights for these sets of variables to allow the estimation of direct and indirect effects of BMI as illustrated in **Supplemental Figures 2C and 2D**.

Comorbidities and the use of supplementary oxygen at home were extracted from medical records and registered as positive if self-reported by patients in a self-administered questionnaire at admission, or if recorded in the preoperative assessment medical note. All participants with OSA self-reported using continuous positive airway pressure (CPAP) during sleep. Altitude was categorized into low (0-1000 m.a.s.l.) and moderate (1000-2500 m.a.s.l.) altitude^22^ due to overspread distribution and inability to model altitude as a non-linear term. Detailed descriptions of all variables are published alongside the dataset elsewhere.^23^

### Statistical analysis

Methods for descriptive analyses and hypothesis testing are detailed in the **Supplemental Methods**. The prevalence of atelectasis with 95% confidence interval (95%CI) was estimated with a one-sample proportion test with Wilson score intervals for the total sample and BMI categories. Due to zero-inflation and skewness, mean atelectasis percentage was determined by bootstrapping with 10,000 re-samples, and 95%CI obtained with the bias-corrected and accelerated (BCa) method. Prevalence ratios (PR) of atelectasis per obesity class (reference category: class 1) were estimated with a modified Poisson regression model with robust errors as described by Yorlets, et al.^24^ and adjusted for age, sex, and altitude. Sensitivity analyses were done by calculating point estimate and lower bound E-values,^25^ which represent the magnitude of association that an unmeasured confounder with the same directionality of effect should have to drive the observed point estimates or lower confidence intervals to the null, respectively.

Atelectasis percentage coverage was modeled in an ordinal logistic regression model.^26^ Despite obesity class not meeting the proportional odds assumption, its impact was checked by comparing against partial proportional odds and multinomial models.^27^ Since the AIC and McFadden adjusted R2 were better for the main proportional odds model,^28^ results of ordinal logistic regression models are presented. Estimates are summarized as the unadjusted and adjusted odds ratio (OR) with Wald 95%CI.

Mean SpO2 was modelled through fractional regression with generalized additive models^29^ with a quasibinomial logit link function to assess the extent to which BMI and atelectasis percentage explained the variation in SpO2. Inverse probability weighting (IPW)^30^ was used to estimate the total effect of BMI on SpO2 in a first model, and the direct (not mediated by atelectasis) and indirect (mediated by atelectasis) effects of BMI on SpO2 in a second model. For the first model, weights (*w1*) were obtained for the exposure-outcome confounders through the non-parametric covariate balancing propensity score (npCBPS) method. For the second model, weights (*w2*) for the mediator-outcome confounders were also obtained with npCBPS and multiplied by *w1*, resulting in overall weights (*w*) used in the model. Since the relationship between exposure-outcome and mediator-outcome were non-linear, the average total, direct, and indirect effects on mean SpO2 value are shown as splines, along with their 95%CI. Goodness-of-fit is shown as the percentage explained deviance (%deviance). Nine influential outliers were removed according to a Cook’s distance greater than 4/n for the two main explanatory variables (BMI n=3, atelectasis percentage n=6); results of analyses keeping all observations can be found elsewhere.^28^

Data were split into SpO2 ≤95% and >95% and modelled separately. In the SpO2 ≤95% subset, the relationships between variables were close to linear, thus allowing estimation of linear effects in a generalized linear model (quasibinomial logit link function), presented as the odds ratio for a change in mean SpO2 for every unit of increase in BMI and atelectasis percentage. The proportion mediated was estimated with the formula 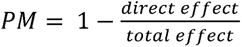 and presented as a percentage. Bootstrap BCa 95%CI for the PM and OR were obtained by re-fitting the models in 10,000 re-samples.

Predicted values of SpO2 for all possible combinations of atelectasis percentage and BMI (within the ranges observed in this study) were obtained from the initial non-linear SpO2 models. The relationship between these three variables was visualized in a three-dimensional plot and predicted curves for every unit-decrease in SpO2 were plotted in a two-dimensional plot. Observed SpO2 values were plotted on top of the gradient of predictions to visualize the range at which combinations are likely to be observed in clinical practice.

Complete-case analysis was performed since missing data was <3% for all variables. Statistical significance was defined as p<0.05. P-values are shown rounded to 3 decimals. All analyses and figures were created with R version 4.3.3. The dataset,^23^ code,^28^ and reports documenting these analyses with references of all R packages used and versions are available at https://github.com/javimangal/preoperative-atelectasis/.

## RESULTS

Out of 281 scheduled surgeries, 35 (12.4%) patients did not present to the hospital, 3 (1.1%) had a positive SARS-CoV-2 antigen test, and the remaining 243 (86.5%) underwent chest CT screening for COVID-19. After exclusion of 7 patients due to CO-RADS ≥3 (n=4) and who reported prior COVID-19 (n=3), 236 were included for analysis.

All participants were residents of the USA and Canada (**Supplemental Figure 3**). The mean age of participants was 40.3 years (SD: 9.87) years and 90.7% were women (n=214). Most patients had a CO-RADS score of 1 (n=230, 97.5%), while the remaining 2.5% (n=6) had CO-RADS 2. Patients with a diagnosis of OSA constituted 14% (n=33) of the sample. The median BMI was 40.3 kg/m^2^ (IQR: 34.6–46.0, range: 30.0–77.3). Most patients were in the class 3 obesity category (n=120, 50.8%), followed by class 1 (n=63, 26.7%) and 2 (n=53, 22.5%). Characteristics of the sample stratified by obesity class are shown in **Table 1**. Pairwise comparisons among independent variables are provided in the reports available in the GitHub repository.^28^

**Table 1.**
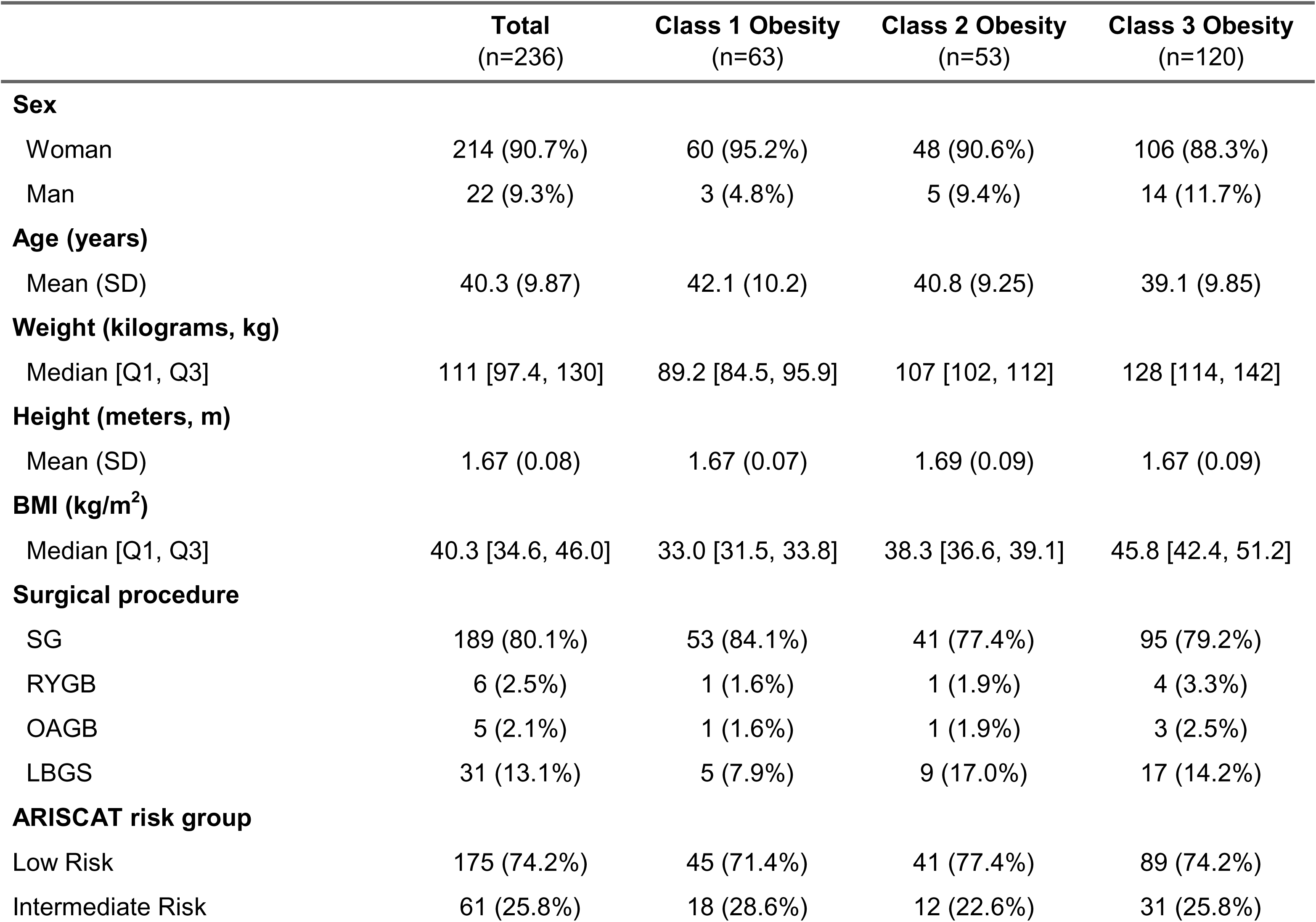
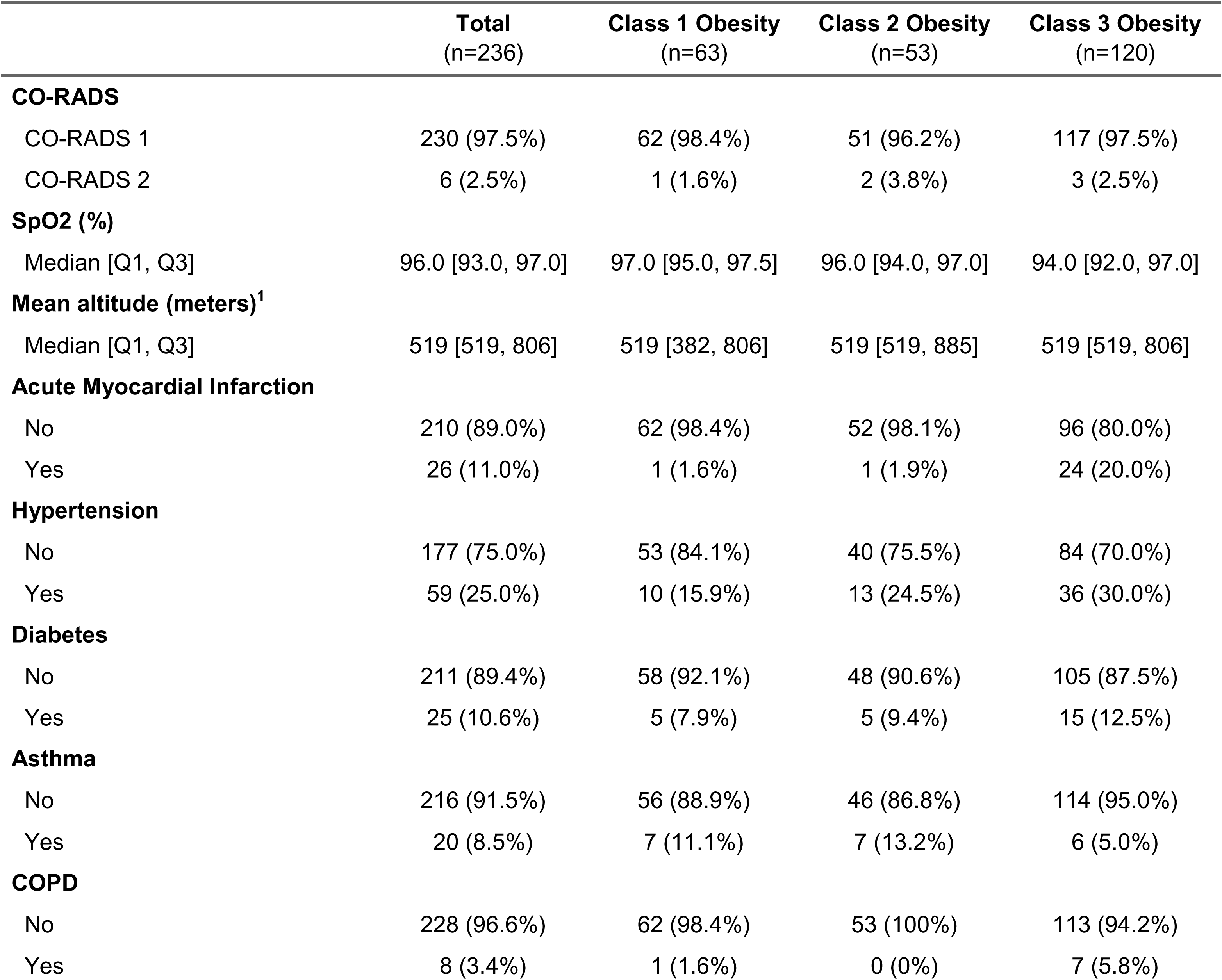
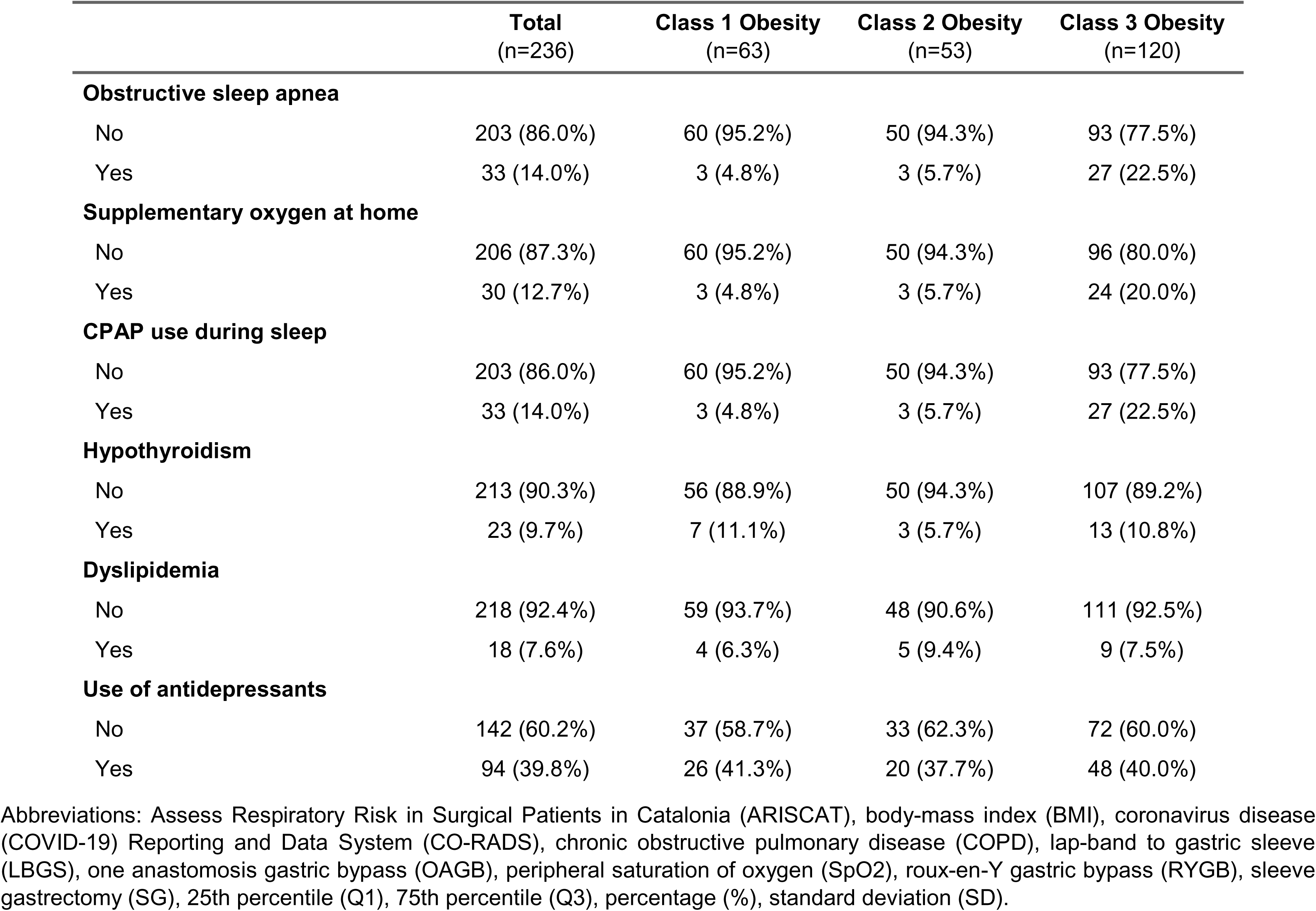
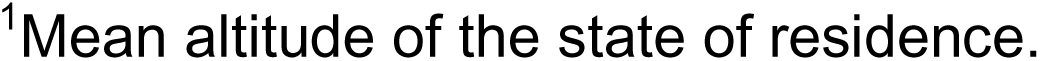
Clinical characteristics of patients, according to WHO obesity categories—class 1 (30-35 kg/m^2^), class 2 (35-40 kg/m^2^), and class 3 (≥40 kg/m^2^) obesity—.

|  | Total<br>(n=236) | Class 1 Obesity<br>(n=63) | Class 2 Obesity<br>(n=53) | Class 3 Obesity<br>(n=120) |
| --- | --- | --- | --- | --- |
| **Sex** |  |  |  |  |
| Woman | 214 (90.7%) | 60 (95.2%) | 48 (90.6%) | 106 (88.3%) |
| Man | 22 (9.3%) | 3 (4.8%) | 5 (9.4%) | 14 (11.7%) |
| **Age (years)** |  |  |  |  |
| Mean (SD) | 40.3 (9.87) | 42.1 (10.2) | 40.8 (9.25) | 39.1 (9.85) |
| **Weight (kilograms, kg)** |  |  |  |  |
| Median [Q1, Q3] | 111 [97.4, 130] | 89.2 [84.5, 95.9] | 107 [102, 112] | 128 [114, 142] |
| **Height (meters, m)** |  |  |  |  |
| Mean (SD) | 1.67 (0.08) | 1.67 (0.07) | 1.69 (0.09) | 1.67 (0.09) |
| **BMI (kg/m<sup>2</sup>)** |  |  |  |  |
| Median [Q1, Q3] | 40.3 [34.6, 46.0] | 33.0 [31.5, 33.8] | 38.3 [36.6, 39.1] | 45.8 [42.4, 51.2] |
| **Surgical procedure** |  |  |  |  |
| SG | 189 (80.1%) | 53 (84.1%) | 41 (77.4%) | 95 (79.2%) |
| RYGB | 6 (2.5%) | 1 (1.6%) | 1 (1.9%) | 4 (3.3%) |
| OAGB | 5 (2.1%) | 1 (1.6%) | 1 (1.9%) | 3 (2.5%) |
| LBGS | 31 (13.1%) | 5 (7.9%) | 9 (17.0%) | 17 (14.2%) |
| **ARISCAT risk group** |  |  |  |  |
| Low Risk | 175 (74.2%) | 45 (71.4%) | 41 (77.4%) | 89 (74.2%) |
| Intermediate Risk | 61 (25.8%) | 18 (28.6%) | 12 (22.6%) | 31 (25.8%) |

|  | **Total**<br>(n=236) | **Class 1 Obesity**<br>(n=63) | **Class 2 Obesity**<br>(n=53) | **Class 3 Obesity**<br>(n=120) |
| --- | --- | --- | --- | --- |
| **CO-RADS** |  |  |  |  |
| CO-RADS 1 | 230 (97.5%) | 62 (98.4%) | 51 (96.2%) | 117 (97.5%) |
| CO-RADS 2 | 6 (2.5%) | 1 (1.6%) | 2 (3.8%) | 3 (2.5%) |
| **SpO2 (%)** |  |  |  |  |
| Median [Q1, Q3] | 96.0 [93.0, 97.0] | 97.0 [95.0, 97.5] | 96.0 [94.0, 97.0] | 94.0 [92.0, 97.0] |
| **Mean altitude (meters)<sup>1</sup>** |  |  |  |  |
| Median [Q1, Q3] | 519 [519, 806] | 519 [382, 806] | 519 [519, 885] | 519 [519, 806] |
| **Acute Myocardial Infarction** |  |  |  |  |
| No | 210 (89.0%) | 62 (98.4%) | 52 (98.1%) | 96 (80.0%) |
| Yes | 26 (11.0%) | 1 (1.6%) | 1 (1.9%) | 24 (20.0%) |
| **Hypertension** |  |  |  |  |
| No | 177 (75.0%) | 53 (84.1%) | 40 (75.5%) | 84 (70.0%) |
| Yes | 59 (25.0%) | 10 (15.9%) | 13 (24.5%) | 36 (30.0%) |
| **Diabetes** |  |  |  |  |
| No | 211 (89.4%) | 58 (92.1%) | 48 (90.6%) | 105 (87.5%) |
| Yes | 25 (10.6%) | 5 (7.9%) | 5 (9.4%) | 15 (12.5%) |
| **Asthma** |  |  |  |  |
| No | 216 (91.5%) | 56 (88.9%) | 46 (86.8%) | 114 (95.0%) |
| Yes | 20 (8.5%) | 7 (11.1%) | 7 (13.2%) | 6 (5.0%) |
| **COPD** |  |  |  |  |
| No | 228 (96.6%) | 62 (98.4%) | 53 (100%) | 113 (94.2%) |
| Yes | 8 (3.4%) | 1 (1.6%) | 0 (0%) | 7 (5.8%) |
| **Obstructive sleep apnea** |  |  |  |  |
| No | 203 (86.0%) | 60 (95.2%) | 50 (94.3%) | 93 (77.5%) |
| Yes | 33 (14.0%) | 3 (4.8%) | 3 (5.7%) | 27 (22.5%) |
| **Supplementary oxygen at home** |  |  |  |  |
| No | 206 (87.3%) | 60 (95.2%) | 50 (94.3%) | 96 (80.0%) |
| Yes | 30 (12.7%) | 3 (4.8%) | 3 (5.7%) | 24 (20.0%) |
| **CPAP use during sleep** |  |  |  |  |
| No | 203 (86.0%) | 60 (95.2%) | 50 (94.3%) | 93 (77.5%) |
| Yes | 33 (14.0%) | 3 (4.8%) | 3 (5.7%) | 27 (22.5%) |
| **Hypothyroidism** |  |  |  |  |
| No | 213 (90.3%) | 56 (88.9%) | 50 (94.3%) | 107 (89.2%) |
| Yes | 23 (9.7%) | 7 (11.1%) | 3 (5.7%) | 13 (10.8%) |
| **Dyslipidemia** |  |  |  |  |
| No | 218 (92.4%) | 59 (93.7%) | 48 (90.6%) | 111 (92.5%) |
| Yes | 18 (7.6%) | 4 (6.3%) | 5 (9.4%) | 9 (7.5%) |
| **Use of antidepressants** |  |  |  |  |
| No | 142 (60.2%) | 37 (58.7%) | 33 (62.3%) | 72 (60.0%) |
| Yes | 94 (39.8%) | 26 (41.3%) | 20 (37.7%) | 48 (40.0%) |

Abbreviations: Assess Respiratory Risk in Surgical Patients in Catalonia (ARISCAT), body-mass index (BMI), coronavirus disease (COVID-19) Reporting and Data System (CO-RADS), chronic obstructive pulmonary disease (COPD), lap-band to gastric sleeve (LBGS), one anastomosis gastric bypass (OAGB), peripheral saturation of oxygen (SpO<sub>2</sub>), roux-en-Y gastric bypass (RYGB), sleeve gastrectomy (SG), 25th percentile (Q1), 75th percentile (Q3), percentage (%), standard deviation (SD).
<sup>1</sup>Mean altitude of the state of residence.

### Preoperative atelectasis

The overall prevalence of preoperative atelectasis was 32.6% (95%CI: 27.0–38.9), being greater in higher obesity classes (p<0.001): class 1 (n=8/63), 12.7% (95%CI: 6.1–24.4); class 2 (n=15/53), 28.3% (95%CI: 17.2–42.6); and class 3 (n=54/120), 45.0% (95%CI: 35.7–53.9). Of those who had atelectasis, right lung base predominance n=53 (68.8%), compared to bilateral lung bases affection n=24 (31.2%). When examining this by obesity class, 87.5%, 66.7%, and 66.7% had right lung base predominance in class 1, 2, and 3 categories:, respectively (p=0.484). Atelectasis percentage showed a non-monotonic non-linear relationship with BMI (**Figure 1A**). A marked increase in atelectasis percentage occurred at BMI higher than ∼45 kg/m^2^. The mean atelectasis percentage coverage in the sample was 2.66% (95%CI:2.07–3.26) and according to WHO categories: class 1 (0.91%, 95%CI:0.32–1.71), class 2 (1.55%, 95%CI:0.75–2.45), and class 3 (4.06%, 95%CI:3.1–5.08). Within class 3 subgroups, the mean atelectasis percentage was 0.7% (95%CI:0.22–1.27) in the 40–45 kg/m^2^ group; 3.64% (95%CI:2.18–5.08), in 45–50 kg/m^2^; and 10.46% (95%CI:8.83–12.5), in the ≥50 kg/m^2^ subgroup. The relative frequencies of the extent of coverage were significantly higher with increasing obesity class (p<0.001) (**Supplemental Figure 4A**), with greater heterogeneity and increasing percentage coverage within class 3 obesity subgroups (**Supplemental Figure 4B**).

**Figure 1.**
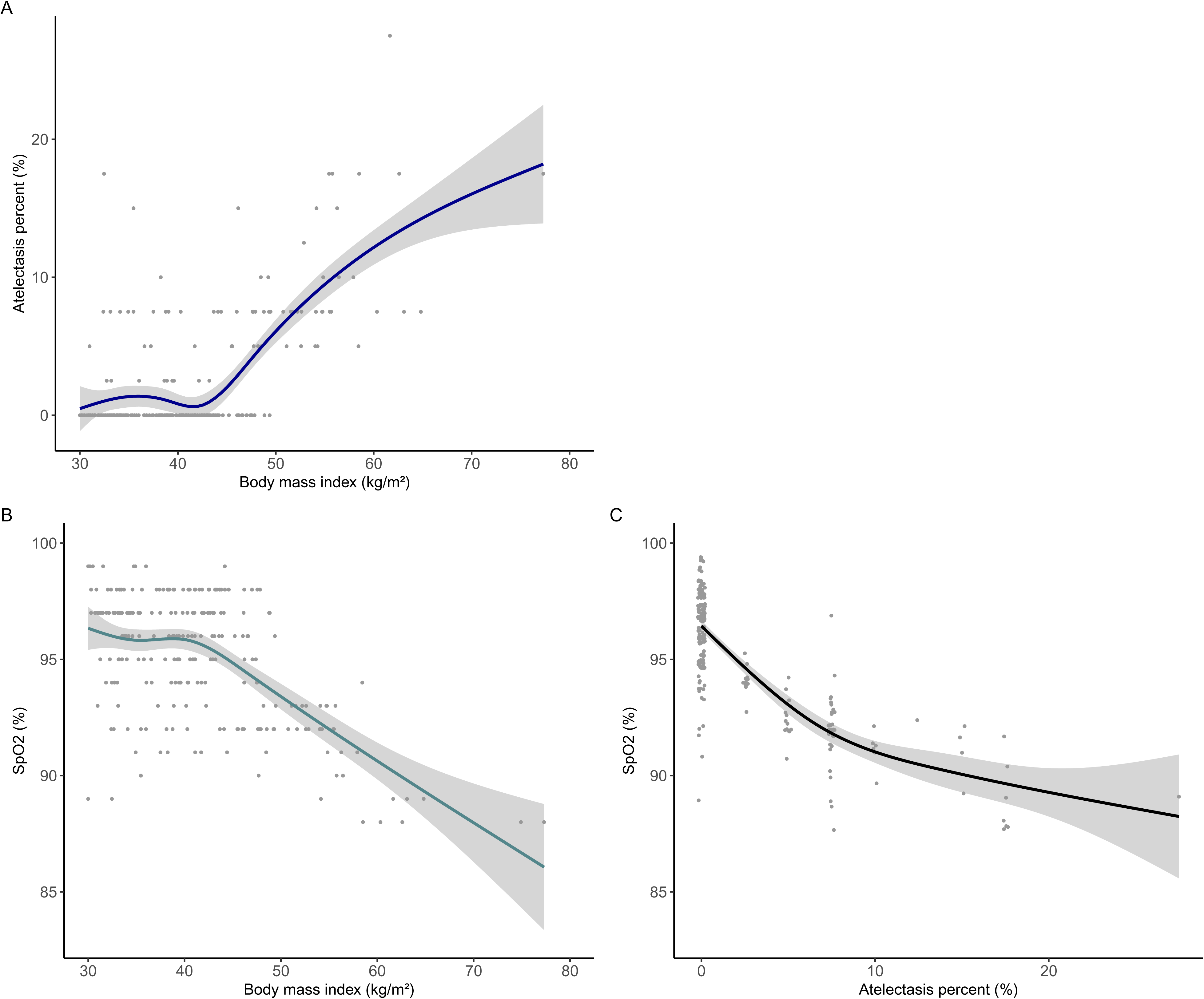
Pairwise non-linear relationships between body mass index, preoperative SpO2, and preoperative atelectasis percentage coverage on chest CT scan. Dots represent individual patient observations. Curves represent the fitted smoothed non-linear relationship. The shaded area corresponds to the 95% confidence interval. **A)** Atelectasis percentage as a function of BMI. **B)** SpO2 as a function of BMI. **C)** SpO2 as a function of atelectasis percentage. Note for design team: We suggest that the empty panel in the upper right corner is used to place the figure legend as we have placed the 3 panels in such a way that y- and x-axes match the same variable.

Age was similarly distributed among patients without atelectasis (40.6, SD:10.1) and those with atelectasis (39.6, SD:9.3) (p=0.498). The differences in atelectasis occurrence between men (45.5%) and women (31.3%) were not statistically significant (p=0.178). Patients with a diagnosis of OSA had atelectasis more frequently (93.9%, n=31/33) than those without (22.7%, n=46/203) (p<0.001). Patients with asthma tended to have atelectasis less frequently (15%) than those without the diagnosis (34.3%) (p=0.079). Unadjusted and adjusted prevalence ratios of atelectasis by obesity class are shown in **Table 2**.

**Table 2.** Crude and adjusted prevalence ratio of atelectasis according to obesity class category.

| Category | PR | 95%CI | aPR<sup>1</sup> | 95%CI | E-value<sup>2</sup> | E-value lower<sup>3</sup> |
| --- | --- | --- | --- | --- | --- | --- |
| Class 1 Obesity |  | Reference |  | Reference |  |  |
| Class 2 Obesity | 2.23 | 1.03—4.84 | 2.17 | 1.0—4.7 | 3.76 | 1.00 |
| Class 3 Obesity | 3.46 | 1.8—6.97 | 3.47 | 1.77—6.83 | 6.40 | 2.94 |
| Subgroups (Class 3 Obesity) |  |  |  |  |  |  |
| 40—45 kg/m<sup>2</sup> | 0.97 | 0.37—2.5 | 0.85 | 0.35—2.1 | 1.63 | - |
| 45—50 kg/m<sup>2</sup> | 3.81 | 1.81—8.01 | 3.52 | 1.63—7.61 | 6.50 | 2.64 |
| ≥50 kg/m<sup>2</sup> | 7.87 | 4.12—15.05 | 8.00 | 4.22—15.17 | 15.48 | 7.91 |

<sup>1</sup>Adjusted for age, sex, and altitude category.
<sup>2</sup>The E-value for unmeasured confounding represents the magnitude of association that an unmeasured confounder with the same directionality of effect should have to drive our point estimate to a value of 1.
<sup>3</sup>E-value for the lower limit of the confidence interval.
Abbreviations: 95% confidence interval (95%CI), adjusted prevalence ratio (aPR), prevalence ratio (PR).

Ordinal logistic regression models were fitted to assess the relationship between increasing obesity class and the extent of atelectasis percentage. The results of univariable and multivariable models are shown in **Table 3**. Compared to class 1 obesity, class 2 obesity was not significantly associated with a greater atelectasis percentage coverage (aOR=2.36, 95%CI: 0.92–6.1), whereas class 3 obesity was associated with a 5-fold increase in the odds of a greater extent of atelectasis percentage (aOR=5.87, 95%CI: 2.57–13.42).

**Table 3.**
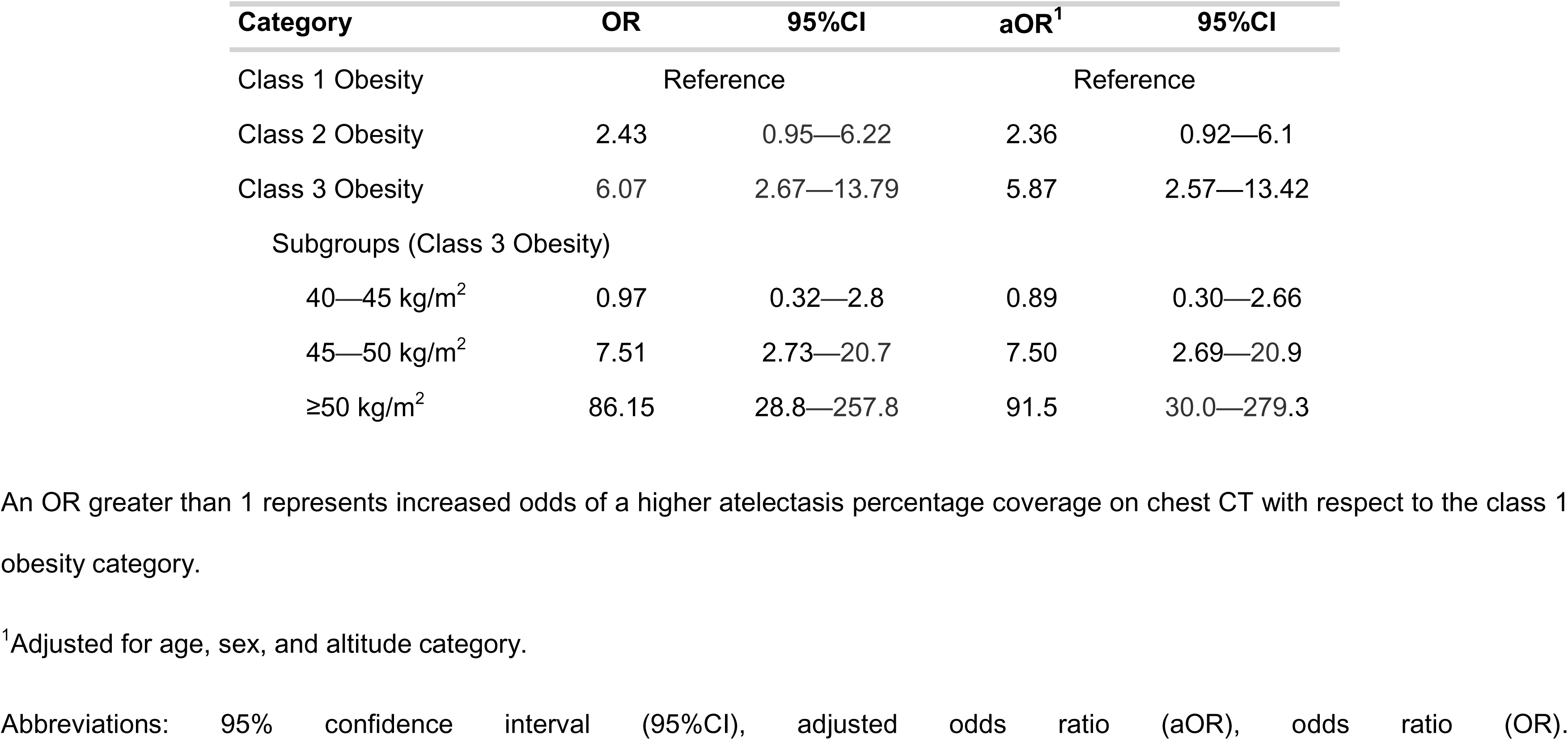
Univariable and multivariable ordinal logistic regression models of lung atelectasis percentage coverage.

| Category | OR | 95%CI | aOR<sup>1</sup> | 95%CI |
| --- | --- | --- | --- | --- |
| Class 1 Obesity | Reference |  | Reference |  |
| Class 2 Obesity | 2.43 | 0.95—6.22 | 2.36 | 0.92—6.1 |
| Class 3 Obesity | 6.07 | 2.67—13.79 | 5.87 | 2.57—13.42 |
| Subgroups (Class 3 Obesity) |  |  |  |  |
| 40—45 kg/m<sup>2</sup> | 0.97 | 0.32—2.8 | 0.89 | 0.30—2.66 |
| 45—50 kg/m<sup>2</sup> | 7.51 | 2.73—20.7 | 7.50 | 2.69—20.9 |
| ≥50 kg/m<sup>2</sup> | 86.15 | 28.8—257.8 | 91.5 | 30.0—279.3 |

An OR greater than 1 represents increased odds of a higher atelectasis percentage coverage on chest CT with respect to the class 1 obesity category.
<sup>1</sup>Adjusted for age, sex, and altitude category.
Abbreviations: 95% confidence interval (95%CI), adjusted odds ratio (aOR), odds ratio (OR).

Due to the heterogeneity observed in atelectasis percentage in the class 3 category, post-hoc analyses were conducted to assess differences in subgroups. The prevalence of atelectasis in the 40–45 kg/m^2^, 45–50 kg/m^2^, and ≥50 kg/m^2^ subgroups was 12.3% (95%CI: 5.5–24.3), 48.4% (95%CI: 30.6–66.6), and 100% (95%CI: 86.7–100), respectively. Prevalence ratios and the OR for an increasing atelectasis percentage coverage are shown in **Table 2** and **Table 3**, respectively.

### SpO2 during the pre-anesthetic assessment

The median SpO2 was 96% (IQR: 93–97), with a minimum value of 88%. A total n=146 (61.9%) had normal SpO2 (above 94%), n=75 (31.8%) had a value in the 90–94% range, and n=15 (6.4%) had ≤90%. BMI exhibited a negative non-linear non-monotonic relationship with SpO2 (**Figure 1B**). SpO2 was significantly lower in patients with atelectasis (92%, IQR: 91–93) compared to those without (97%, IQR: 96–98) (p<0.001), and lower in patients with bilateral lung base atelectasis (91.5%, IQR: 90–92) compared to those with right lung predominance (92%, IQR: 92–93) (p=0.006). There was a decreasing trend in SpO2 with higher atelectasis percentage extension (**Figure 1C**). Patients with OSA had a lower median SpO2 (92%, IQR: 91–93) than those without (96%, IQR: 94–97) (p<0.001). SpO2 was not correlated (rho= -0.065, p=0.32) with the values of hemoglobin (mean:14.5, SD:1.21 g/dL) observed in this study. Similarly, mean altitude of the place of residence (range: 31–1861 m.a.s.l.), age (rho= 0.022, p=0.74), and sex (p=0.413) were not significantly associated with SpO2.

The total effect of BMI on the mean SpO2 across different values of BMI is shown in **Figure 2A**, while the total effect of atelectasis coverage on chest CT on SpO2 is shown in **Figure 2C**. In the IPW model including both terms, the direct effect of BMI on SpO2 was significantly reduced to null when controlled for atelectasis percent (**Figure 2B**), whereas the effect mediated trough atelectasis (indirect effect of BMI) remained largely unchanged when compared to that not adjusted for BMI (**Figure 2D**), thus indicating that the effect of BMI on SpO2 was largely mediated by atelectasis. Predicted SpO2 values from this model are shown in **Figure 3A** and plotted alongside observed SpO2 values (**Figure 3B**). The 3-dimensional relationship between SpO2 predictions, BMI and atelectasis percentage is shown in **Supplemental Figure 5**.

**Figure 2.**
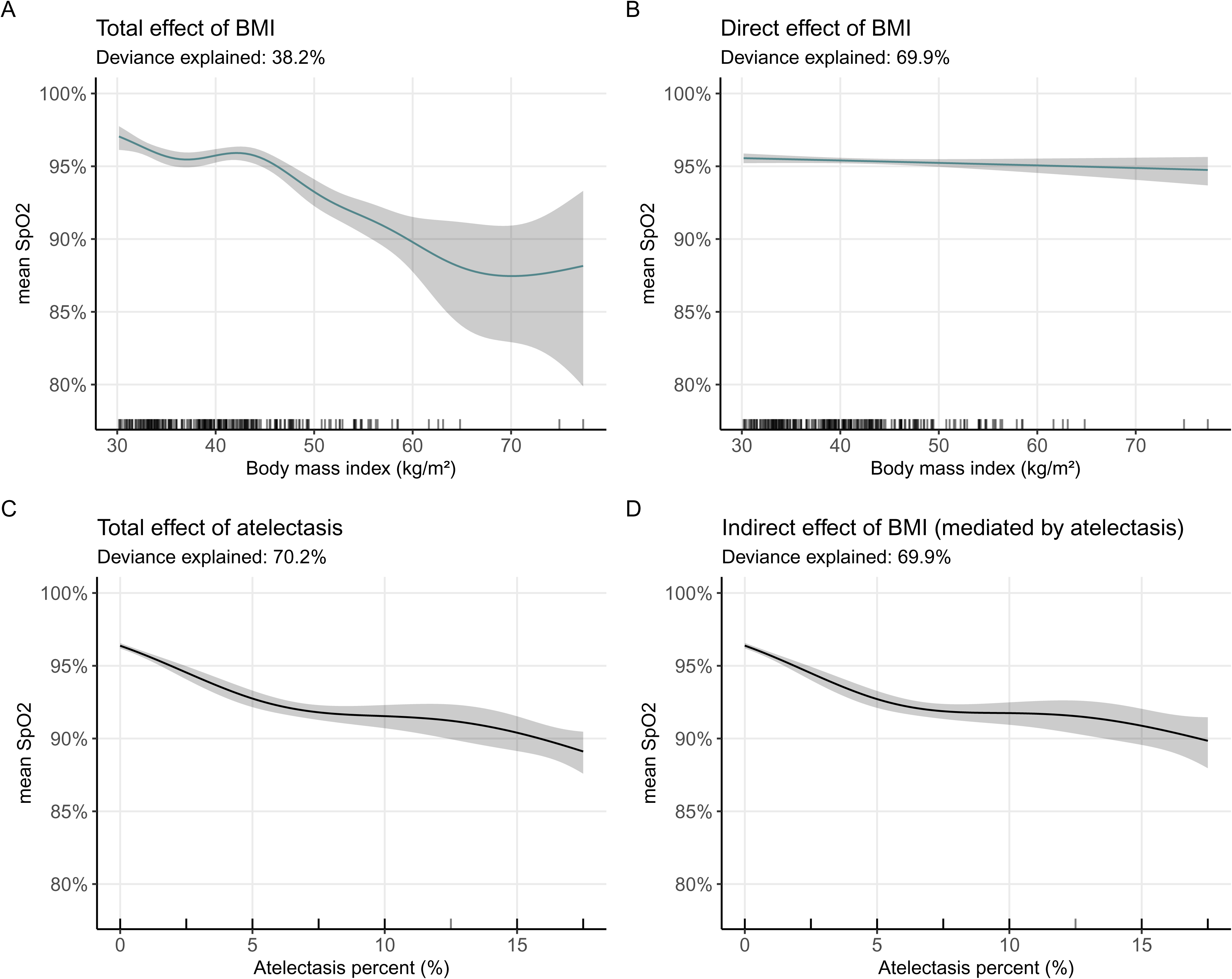
Total, direct (not mediated), and indirect (mediated through atelectasis) effects of body mass index (BMI) on mean SpO2 during the preoperative assessment. Solid lines represent the partial effect on mean SpO2 for increasing BMI (blue green) and atelectasis percent coverage on chest CT (black), with 95% confidence intervals (shaded area). Inverse probability weighted* models were used to present **A)** Total effect of BMI in a model including only a smooth term for BMI (p<0.001). **B)** Direct effect of BMI (not mediated by atelectasis), with a smooth term for BMI (p=0.182) controlled for the effect of atelectasis. **C)** Total effect of atelectasis percent coverage on chest CT in a model including only a smooth term for atelectasis percent (p<0.001). **D)** Indirect effect of BMI (mediated by atelectasis), with a smooth term for atelectasis percent (p<0.001) controlled for the effect of BMI. *Inverse probability weights were obtained for the following set of variables: - *w1* (age, sex, and altitude), panel A; - *w2* (BMI, age, sex, altitude, obstructive sleep apnea, asthma, and COPD), panel C; - *w* (product of *w1* and *w2*), panels B and D.

**Figure 3.**
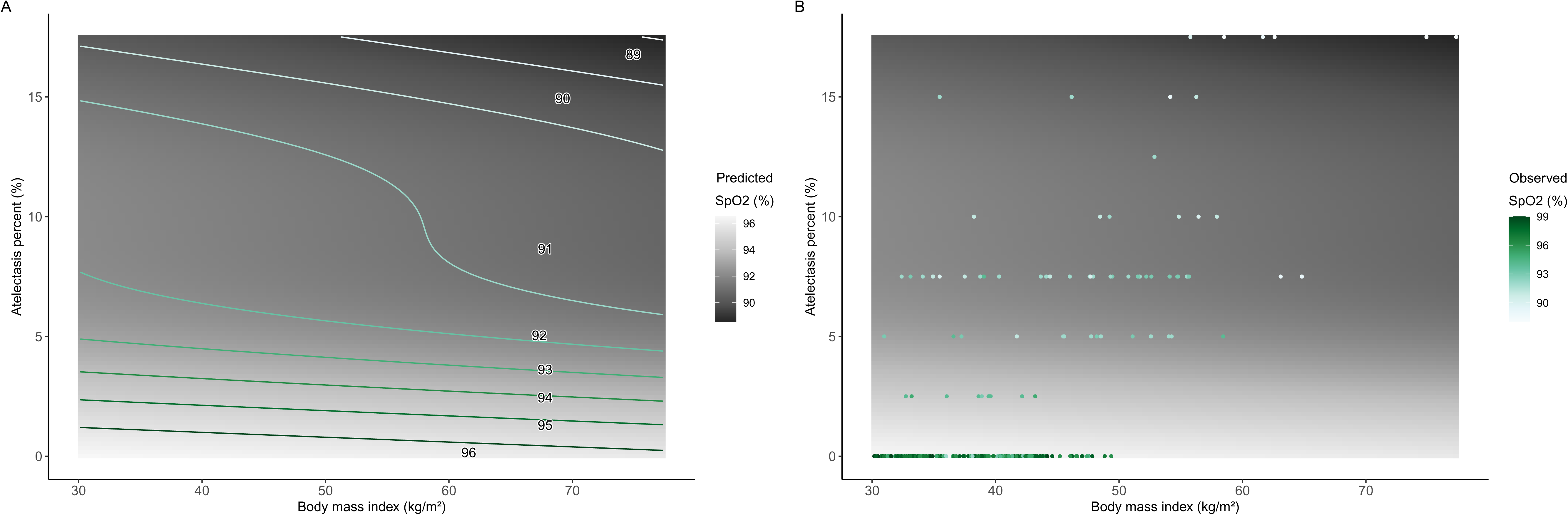
Predicted and measured SpO2 values by body mass index (BMI) and atelectasis percentage on chest CT scan. **A)** Predicted SpO2. Curved lines correspond to SpO2 values that are predicted by BMI and atelectasis percentage. The background grid on a gray scale corresponds to predicted values of SpO2 for every possible combination of atelectasis percentage and BMI, weighted for the set of variables *w* (see legend in Figure 2 for explanation). **B)** Observed SpO2. Every dot corresponds to an individual patient observation located at the exact BMI and atelectasis percentage measured, colored according to the observed SpO2 value.

Analysis of residuals suggested that the model explained values of SpO2 ≤95%, but not those >95%, reason why data were split and modelled separately. All participants but one in the >95% dataset had atelectasis, reason why modelling was not possible in this subset. In the ≤95% group, relationships between variables were close to linear and the OR for the change in mean SpO2 for every unit increase in BMI and atelectasis percent are provided in **Supplemental Table 1**. The proportion of the effect of BMI on SpO2 mediated through atelectasis was 81.5% (95%CI: 56.0–100).

## DISCUSSION

Our results show a high prevalence of atelectasis in obese patients before surgery, with prevalence increasing with higher BMI. An increased risk of higher atelectasis compared to the class 1 obesity category was only confidently estimated at BMI ≥45 kg/m^2^. Since our study only included patients with a BMI ≥30 kg/m^2^, it is likely that these prevalence ratios would be even higher if overweight or normal BMI were set as the reference categories, as prior studies show that atelectasis occur more frequently before surgery in obese patients than those with normal BMI.^8^ Thus, our results only allow us to conclude an increased relative risk of atelectasis at BMI ≥45 kg/m^2^ compared to obese patients in the lowest obesity category (BMI 30-35 kg/m^2^).

Although the prevalence of atelectasis in our study may seem high compared to other studies reporting low proportions of atelectasis even during the postoperative period—for instance, 0.4% in the BOLD registry^31^ and 4.4–5.6% in the PROBESE trial^32^—, atelectasis assessment in such studies was based on reporting of atelectasis after indication of an imaging study upon clinical suspicion. Therefore, patients in whom imaging is not performed are assumed to not have atelectasis, which biases the estimate towards the null. Here, we propose a definition of atelectasis (≥2.5% of atelectasis coverage on chest CT as a fraction of total lung volume) which could be used in future prospective studies to homogenize outcome assessment and reporting of atelectasis.

We found a mean overall atelectasis percentage coverage (as a fraction of total lung volume) of 2.66% (95%CI:2.07–3.26), which is close to the 2.1% reported by Eichenberger, et al.^8^ Lower numbers (0.4 ± 0.7%) were reported by Reinius, et al.,^15^ although their measurement was at the end of expiration and their estimate could be biased to the null due to zero-inflation as suggested by the SD which includes negative values. Atelectasis percentage in our study increased at higher BMI: class 1 (0.91%), class 2 (1.55%), and class 3 (4.06%).

When modelling BMI as a continuous variable, there was a marked increase in atelectasis percentage at a cutoff close to 45 kg/m^2^. Ordinal logistic regression analyses showed that the odds of having a higher atelectasis percentage coverage is higher in 5 to 6 orders of magnitude in the class 3 obesity category, but not statistically significant in the class 2 obesity subgroup despite the point estimate being greater than 1 (aOR=2.36, 95%CI:0.92–6.1). We observed that only categories above 45 kg/m^2^ had a significantly higher odds of increased atelectasis percentage coverage on chest CT after adjusting for confounders. The reason why the prevalence of atelectasis and atelectasis percentage coverage increased in class 2 obesity and then decreased again to levels comparable to class 1 obesity in the 40-45 kg/m^2^ subgroups could be due to outliers, random variation, and the latent possibility of residual confounding driving transient increases. Sensitivity analysis with E-values present the magnitude of association that a latent confounder with the same directionality of effect should have to explain these associations, showing that our results for BMI >45 kg/m^2^ are strong estimates.

Besides the mechanisms discussed in the introduction, these findings could also be related to the significant reduction in functional residual capacity and dynamic compliance (Cdyn) due to increasing intraabdominal and intrathoracic pressure with higher BMI. This phenomenon has been shown by Steier et al.^33^ when comparing normal-BMI to obese patients (0.135 L/cmH2O vs 0.105 L/cmH2O, p<0.05) and Li et al.^34^ (∼0.031 L/cmH_2_O after intubation).

As a secondary objective, we studied the extent to which SpO2 measured during the preoperative assessment could be due to BMI alone or mediated by atelectasis percentage. Strikingly, the plots of atelectasis percent vs BMI (Figure 1A) and SpO2 vs BMI (Figure 1B) are nearly mirror-images. We showed that atelectasis alone explained 70.2% of variation in SpO2 and that the effect of BMI was largely mediated by atelectasis since the effect of BMI on SpO2 was nearly completely attenuated when adjusting for atelectasis percentage, whereas the opposite was not true. Moreover, in the subset of participants with SpO2 <96% where the relationships were close to linear, the proportion of the effect mediated by atelectasis was 81.5% (95%CI: 56.0–100).

Showing that atelectasis in the supine position can explain SpO2 in the seated position challenges the notion that atelectasis in lung-dependent regions are merely postural with often little clinical relevance,^35^ or artifacts expected to solve by performing chest CT in the prone position^36^. Furthermore, the presence of atelectasis with consequences in SpO2 of obese patients before elective surgery strengthens our understanding of the pathophysiology of obesity, possibly implying that chronic hypoxemia in patients with obesity could be largely explained by mechanisms leading to atelectasis^37^ and potentially related to development of chronic dyspnea^38^, intolerance to exercise^39^, or pulmonary hypertension through hypoxic vasoconstriction^40^.

The predictions plots showed that only the curves for SpO2 values between 89-96% are well represented. While it would not be possible to extend the y-axis to values lower than 0% to predict SpO2 >96%, it would be possible to extend the y-axis to higher atelectasis percentage values, meaning that SpO2 values lower than 88% could likely be predicted. We found these results encouraging since they show that predicting preoperative atelectasis without the need of performing a chest CT is possible and something that could be implemented in future studies. As this is a proof-of-concept, we advise against immediate implementation and we instead encourage researchers to develop and validate a prediction model of atelectasis percentage coverage on large-scale, sufficiently powered, and representative studies.^41^ Prediction models of preoperative atelectasis could be used to plan and guide optimal intraoperative ventilation parameters since individualized ventilation strategies have shown to be promising for preventing postoperative atelectasis,^42^ while also allowing to estimate the extent to which postoperative atelectasis are clinically relevant with respect to the extent of atelectasis that were already present before surgery.

Strengths of our study include the large sample of patients with obesity, including extreme BMI, with a high inclusion rate of 84% of all programmed surgeries. Additionally, the availability of chest CT scans before surgery due to the COVID-19 pandemic that allowed us to study the prevalence of preoperative atelectasis. Furthermore, we implemented DAG-informed modelling which is currently the recommended approach to study potentially causal relationships.^43^

Limitations of our study include that atelectasis percentage was rounded to the lowest 2.5% category, which caused loss of information. This could have led to an underestimation of the prevalence of atelectasis. Thus, our results are likely conservative, which is pertinent for a preliminary study. Additionally, we did not have good documentation in medical records of other potential confounders like recent respiratory infections or heart disease. Although COVID-19 could be an additional potential confounder, we excluded participants who declared a prior history of COVID-19 or who had suggestive findings of current SARS-CoV-2 infection according to CO-RADS. One additional limitation is that our study is poorly representative of men since 90.7% were women, although this is a common situation in studies conducted in bariatric surgery (70-79% women).^8,15,44,45^ Furthermore, our sample was overall younger than other large representative studies which could explain why the prevalence of comorbidities like hypertension and diabetes were lower.^45^ Lastly, our study was restricted to the preoperative period due to limited resources, reason why future studies could attempt to explore preoperative atelectasis and their impact in postoperative outcomes.

## CONCLUSION

The overall prevalence of preoperative atelectasis in patients with obesity undergoing bariatric surgery was 32.63% (95%CI: 26.97–38.85) and increased with higher obesity categories. The mean atelectasis percentage coverage in chest CT was 2.66% (95%CI:2.07–3.26) and similarly increased with higher BMI. The risk of having a greater prevalence and extension of atelectasis were significantly higher at BMI ≥45 kg/m^2^. Preoperative atelectasis mediated the effect of BMI on preoperative SpO2, with atelectasis alone explaining 70% of the variability in SpO2.

## Supporting information

Supplemental Content

## Data Availability

The data that support the findings of this study are openly available in the Harvard Dataverse at https://doi.org/10.7910/DVN/4JZZLB. The code documenting the analysis in this study is openly available in https://github.com/javimangal/preoperative-atelectasis/ and deposited in https://zenodo.org/doi/10.5281/zenodo.10211241.

https://github.com/javimangal/preoperative-atelectasis/

https://zenodo.org/doi/10.5281/zenodo.10211241

