## Supplemental Content for "Preoperative atelectasis in patients with obesity undergoing bariatric surgery: a cross-sectional study"

### Supplemental Contents

### Supplemental Methods

#### *Coronavirus disease (COVID-19) screening*

During the study period, a joint initiative between Mexico, Canada and the United States (USA) restricted international non-essential travels due to the COVID-19 emergency,[1] but travelling for elective surgeries was possible. American Society of Anesthesiologists (ASA) recommendations for elective surgeries included that patients were tested against SARS-CoV-2, screened for symptoms of COVID-19, and advised against surgery when symptoms were present.[2] Rapid antigen tests against SARS-CoV-2 were not yet available in Mexico[3] and it was not feasible to perform RT-PCR on patients before surgery due to long waiting times until result reporting. Thus, the hospital committee decided that patients were screened for COVID-19 by a sequential approach consisting of 1) patients were asked for sign and symptoms of COVID-19 prior to arriving to the hospital and advised not to present for surgery if these were present, 2) upon arrival at the hospital, rapid SARS-CoV-2 antibody testing was performed. If IgM against SARS-CoV-2 was positive, the surgery was postponed. 3) If the antibody test was negative, a chest computed tomography (CT) was performed and a CO-RADS[4] score ≥3 was considered suggestive of COVID-19, leading to cancellation of the surgery.

Since chest CT images were available as part of this screening process, we considered this a unique opportunity to study the prevalence and extent of preoperative atelectasis in patients with obesity undergoing bariatric surgery.

***Sample size estimation***

We did not identify prior studies reporting the prevalence of preoperative atelectasis in patients with obesity. Thus, we calculated the minimum sample size under the following assumptions:

1. The PROBESE trial reported that 5.6% of patients in the high PEEP group (12 $cmH_2O$) and 4.4% in the low PEEP group (4 $cmH_2O$) had postoperative atelectasis.[5] Assuming a linear relationship between PEEP and atelectasis, patients with no PEEP (0 $cmH_2O$) could be expected to have a prevalence of atelectasis of 6%.
2. A deviation of 5% in the expected prevalence estimate was selected according to recommendations by Naing et al.[6] for preliminary small-scale studies.
3. Confidence level of 95%.

The minimum sample size obtained was 241 patients.

***Relationships between variables – directed acyclic graph (DAG)***

Supplemental Figure 1 presents the main relationships between exposure $\rightarrow$ outcome with atelectasis as the main outcome of interest, whereas Supplemental Figure 2 shows relationships relevant to mediations analyses, including the rationale and assumptions for inverse probability weighting. The following subsections explain the specific evidence or reasons used to build these DAGs.

**Exposure**

The increasing degree of obesity, according to the WHO obesity class categories or BMI, is the exposure of interest and is represented with a green node (*type_obesity*).

**Primary outcome**

Having atelectasis (Yes or No) in the prevalence ratio models, and an increasing degree of atelectasis (atelectasis percent) in the ordinal regression models are the main outcomes of interest. A green arrow from *type_obesity* to *atelectasis_percent* represents the exposure-outcome relationship of interest in the DAG.

**Secondary outcome**

Decreasing preoperative SpO2 is hypothesized to be related to an increasing degree of obesity. An arrow from type_obesity to spo2_VPO at the upper part of the figure represents this. Atelectasis_percentage is thought to be the main mediator of the effect of BMI on preoperative SpO2. An arrow from type_obesity to atelectasis_percent, followed by an arrow from atelectasis_percent to spo2_VPO.

**Covariates**

*Sex and Age*

These two variables are known to be associated with a higher risk of developing postoperative atelectasis in patients with obesity undergoing bariatric surgery.[7] Arrows originating from these variables and going to type_obesity, atelectasis_percent, and spo2_VPO represent these relationships.

The implications for statistical analyses is that ***sex*** and ***age*** are both **confounders** to be accounted for in both of the models with atelectasis and SpO2 as outcomes.

*Obstructive sleep apnea*

Increasing BMI is a strong risk factor for OSA and OSA severity.[7] In a directed acyclic graph, all arrows should have a single direction. Since obesity is the risk factor leading to development of obstructive sleep apnea, and not the other way around,[8] an arrow originating in type_obesity, pointing towards OSA represents this relationship. Thus, OSA is hypothesized to lead to the degree of atelectasis and preoperative SpO2. Therefore, an arrow from OSA to atelectasis_percent and spo2_VPO represents these relationships. The implications for the analysis are the following:

1. OSA is a potential **mediator** of the effect of BMI on atelectasis percentage. Therefore, this variable should **not** be adjusted for in the models with ***atelectasis*** as the outcome.
2. OSA is a **confounder** of the mediator-outcome relationship in the models with ***SpO2*** as the outcome.

*Asthma*

It has been shown that obesity leads to asthma, whereas the inverse relationship is very unlikely to be possible.[9] Thus, an arrow from type obesity to asthma was drawn.

It has been reported that obesity-associated late onset non-allergic asthma is negatively related to atelectasis due to a tendency to develop more air trapping than atelectasis in these patients, compared to patients with obesity and no diagnosis of asthma in whom the airways collapse slowly and air is expelled, leading to atelectasis.[10] During sleep, asthma affects SpO2 independently of BMI and OSA.[11] For these reasons, an arrow from asthma to atelectasis, and an arrow from asthma to SpO2 was drawn.

The implications for the analysis are the following:

1. Asthma is a potential **mediator** of the effect of BMI on atelectasis percentage. Therefore, this variable should **not** be adjusted for in the models with ***atelectasis*** as the outcome.
2. Asthma is a potential **confounder** of the mediator-outcome relationship in the models with ***SpO2*** as the outcome.

*Chronic obstructive pulmonary disease (COPD)*

Although there is a strong relationship between undernutrition and COPD, the relationship between obesity and COPD has been inconsistent among studies. Since this study only included patients with obesity, the potential relationship between underweight and COPD is likely not relevant for this study. Furthermore, there is still doubt regarding any potential role of obesity-related pathophysiological mechanisms which could potentially lead to COPD.[12] For these reasons, an arrow between COPD and obesity was not drawn. This assumption was checked through the conditional independencies checking method (see Part 4 report[13]), thus confirming that this assumption is consistent with the data.

Regarding a relationship between COPD and SpO2, there is a clear relationship between these variables, reason why an arrow going from COPD to SpO2 was drawn. As for atelectasis, studies have found atelectasis is related with COPD, especially in patients with wood smoke-related COPD.[14,15] Thus, an arrow from COPD to atelectasis was drawn.

The implications for the analysis are the following:

1. COPD is **not** a potential confounder of the effect of BMI on atelectasis percentage. Therefore, this variable should **not** be adjusted for in the models with ***atelectasis*** as the outcome.
2. COPD is a potential **confounder** of the mediator-outcome relationship in the models with ***SpO2*** as the outcome.

*Altitude*

Although not directly linked to obesity, participants with OSA at an altitude above 1600 meters can develop hypobaric hypoxia, which "promotes frequent central apneas in addition to obstructive events, resulting in combined intermittent and sustained hypoxia".[16]

For the atelectasis outcome, we did not find evidence either supporting or rejecting an association between altitude and prevalence of atelectasis. However, during the conditional independencies assumptions testing procedure, the data suggested a correlation, reason why an arrow from altitude to atelectasis was drawn as the reverse is less likely to be true (i.e., obesity would hardly determine the altitude of the place of residence).

The implication for analysis is that ***altitude_cat*** is a potential **confounder** to be accounted for in both models (with atelectasis and SpO2 as outcomes).

*Oxygen use and continuous positive airway pressure (CPAP) use at home*

These variables are **colliders** and **descendants** of the exposure, mediator, outcomes, and covariates of interest. The implications for analyses are that these 2 variables should **not** be adjusted for in any of the models as this would introduce **collider bias**.

*Hemoglobin*

There is no strong evidence supporting a link between BMI and hemoglobin. In any case, hemoglobin would be a descendant of all main variables of interest (exposure, mediator, and outcomes). Thus, hemoglobin was excluded from this DAG for simplification.

*Other variables*

Other variables that are potential confounders are not shown in this DAG since they were addressed by design in this study as follows:

- Current COVID-19: Exclusion criteria were applied to **n=2** patients with CO-RADS 3 and **n=2** with CO-RADS 4. Only participants with low probability of COVID-19 (CO-RADS 1 and 2) were included in this study.
- Prior COVID-19: This was an exclusion criterion (**n=3**).
- Bronchiectasis in chest CT: This was an exclusion criterion (n=0).
- Neuromuscular diseases: This was an exclusion criterion (n=0).
- Prior or current history of tuberculosis: This was an exclusion criterion (n=0).

*Unmeasured variables*

Due to the possibility of unmeasured confounders, E-values[17] were calculated and presented when possible as sensitivity analyses.

***Statistical analysis***

Descriptive characteristics are presented as the mean and standard deviation (SD), median with interquartile range (IQR), and/or range (minimum–maximum) for numerical variables. For categorical variables, absolute frequencies and percentages were calculated. Categorical variables were summarized in frequency tables and compared with the chi-squared test. Relationships between 2-level categorical and numerical variables were assessed through stacked histograms, boxplots, and Q-Q plots and compared with unpaired t-tests or Wilcoxon's test. Scatterplots were used to assess relationships between numeric variables and compared with Spearman's rank test if monotonically increasing/decreasing.

When relationships were non-linear, an optimal smooth curvature was determined in a general additive model by increasing the number of knots and comparing against linear terms; if the non-linear term was significantly better than linear, the lowest k with a drop ≥2 in the Akaike information criterion (AIC), plus an adequate visual representation of the relationship and optimal k-index was selected.

A map of the USA and Canada territories was plotted showing the absolute frequency of patients per state of residence. Relative frequencies of atelectasis percentage by obesity category and class 3 subgroups (40–45, 45–50, and ≥50) are shown in a barplot.

***References***

1. U.S. Department of Homeland Security. DHS Measures on the Border to Limit the Further Spread of Coronavirus. Fact Sheets 2020. Available at: https://www.dhs.gov/news/2020/05/20/fact-sheet-dhs-measures-border-limit-further-spread-coronavirus. Accessed June 16, 2020.
2. American Society of Anesthesiologists. ASA/APSF Statement on Perioperative Testing for the COVID-19 Virus Patients. COVID-19 and Elective Surgery 2020:1–3. Available at: https://www.asahq.org/in-the-spotlight/coronavirus-covid-19-information/elective-surgery. Accessed June 3, 2020.
3. Mancilla-Galindo J, Kammar-García A, Martínez-Esteban A, Meza-Comparán HD, Mancilla-Ramírez J, Galindo-Sevilla N. COVID-19 patients with increasing age experience differential time to initial medical care and severity of symptoms. Epidemiology and Infection 2021;149:e230.
4. Prokop M, Everdingen W van, Rees Vellinga T van, Quarles van Ufford H, Stöger L, Beenen L, Geurts B, Gietema H, Krdzalic J, Schaefer-Prokop C, Ginneken B van, Brink M, COVID-19 Standardized Reporting Working Group of the Dutch Radiological Society. CO-RADS: A Categorical CT Assessment Scheme for Patients Suspected of Having COVID-19-Definition and Evaluation. Radiology 2020;296:E97–104.
5. Bluth T, Serpa Neto A, Schultz MJ, Pelosi P, Gama de Abreu M, Bobek I, Canet JC, Cinella G, Baerdemaeker L de, Gregoretti C, Hedenstierna G, Hemmes SNT, Hiesmayr M, Hollmann MW, Jaber S, Laffey J, Licker MJ, Markstaller K, Matot I, Mills GH, Mulier JP, Putensen C, Rossaint R, Schmitt J, Senturk M, Severgnini P, Sprung J, Vidal Melo MF, Wrigge H, Abelha F, et al. Effect of Intraoperative High Positive End-Expiratory Pressure (PEEP) with Recruitment Maneuvers vs Low PEEP on Postoperative Pulmonary Complications in Obese Patients: A Randomized Clinical Trial. JAMA - Journal of the American Medical Association 2019;321:2292–305.
6. Naing L, Nordin R Bin, Abdul Rahman H, Naing YT. Sample size calculation for

prevalence studies using Scalex and ScalaR calculators. BMC Medical Research Methodology 2022;22:209.

13. Mancilla-Galindo J. Preoperative Atelectasis in Bariatric Surgery. 2023.

14. González-García M, Gomez DM, Torres-Duque CA, Barrero M, Villegas CJ, Pérez JM, Varon H. Tomographic and functional findings in severe COPD: comparison between the wood smoke-related and smoking-related disease. Jornal Brasileiro de Pneumologia 2013;39:147–54.

15. Moreira MAC, Barbosa MA, Queiroz MC de CAM de, Teixeira KISS, Torres PPT e S, Santana Júnior PJ de, Montadon Júnior ME, Jardim JR. Pulmonary changes on

HRCT scans in nonsmoking females with COPD due to wood smoke exposure. Jornal Brasileiro de Pneumologia 2013;39:155–63.

16. Bloch KE, Latshang TD, Ulrich S. Patients with Obstructive Sleep Apnea at Altitude. High Altitude Medicine & Biology 2015;16:110–6.

17. VanderWeele TJ, Ding P. Sensitivity Analysis in Observational Research: Introducing the E-Value. Annals of Internal Medicine 2017;167:268.

**Supplemental Figure 1**. Directed acyclic graph (DAG) for the primary outcome.

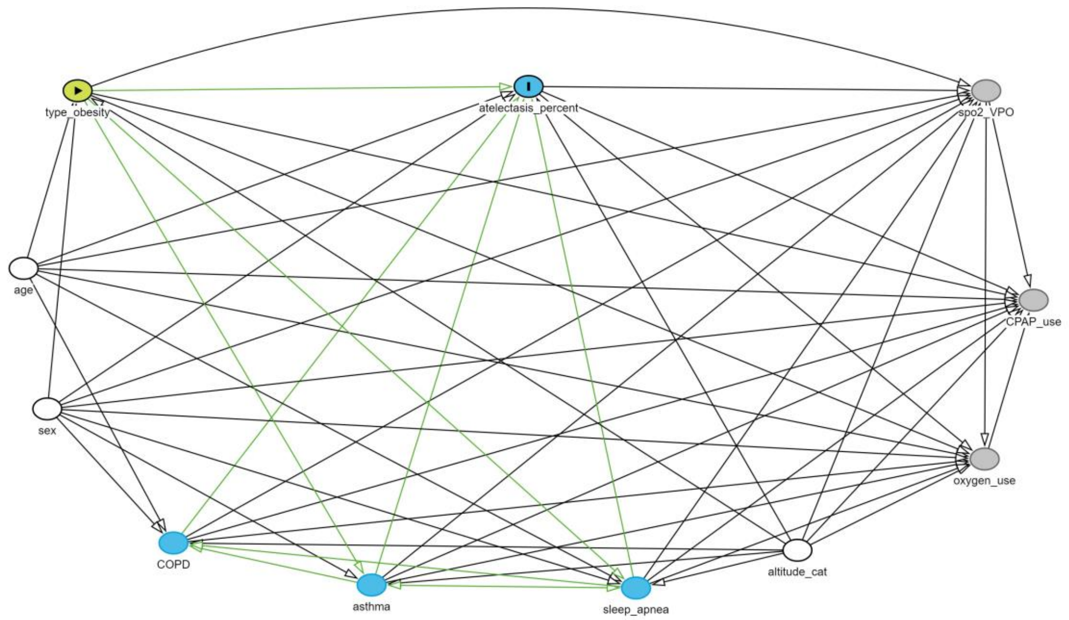

Each circle (node) represents a variable. The green node with an arrowhead inside is the exposure of interest (obesity class category), while the node filled in blue with an "I" inside is the main outcome of interest (atelectasis percent). White nodes (age, sex, and altitude category) correspond to the minimal set of confounders to adjust for to remove biasing pathways. Blue nodes (COPD, asthma, and obstructive sleep apnea) are potential mediators of the effect that should not be adjusted for in the models with atelectasis as the outcome. The gray nodes (SpO2, CPAP use at home, and supplementary oxygen use at home) are descendants and colliders, which should not be adjusted for in the models with atelectasis as the outcome. Black arrows show the direction of association between variables, while green arrows are potential causal paths between exposure and outcome.

This DAG and its associated model code can be accessed at: https://dagitty.net/mhiUeH9YK.

**Supplemental Figure 2***.* Directed acyclic graphs (DAGs) for the secondary outcome and mediation analysis with inverse probability weighting.

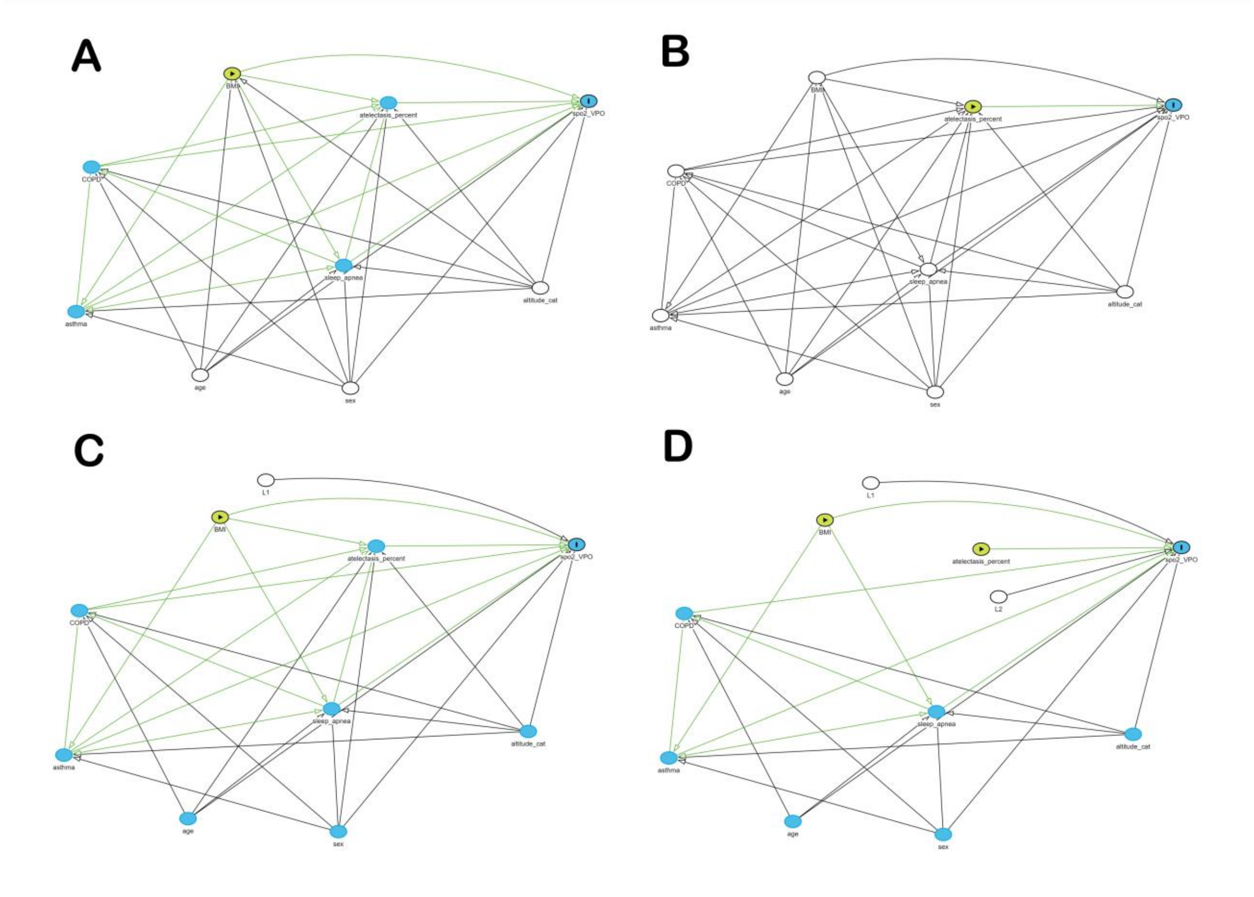

Relationships between variables relevant to determine the effects on SpO2 (outcome). **A)** To determine the total effect of BMI, the minimal adjustment set *L1* consists of age, sex, and altitude, whereas **B)** the total effect of atelectasis percentage requires adjustment for the set *L2* (BMI, age, sex, altitude, obstructive sleep apnea, asthma, and COPD). Through inverse probability weighting, **C)** the total effect of BMI can be modelled by first obtaining weights (*w1*) for the *L1* set, and **D)** the total effect of atelectasis percentage can similarly be obtained by modelling with weights (*w2*) for the *L2* set. Finally, the direct effect of BMI (not mediated by atelectasis) and the indirect effect of BMI (mediated through atelectasis) can be modelled by first obtaining combined weights (*w* = *w1***w2*) and including both variables in the model.

**Supplemental Figure 3.** State of residence of participants.

| State | n |
|---|---|
| Texas | 81 |
| Washington | 39 |
| Utah | 27 |
| Alberta | 21 |
| Florida | 20 |
| California | 16 |
| Louisiana | 16 |
| Arkansas | 7 |
| Missouri | 5 |
| Nevada | 4 |

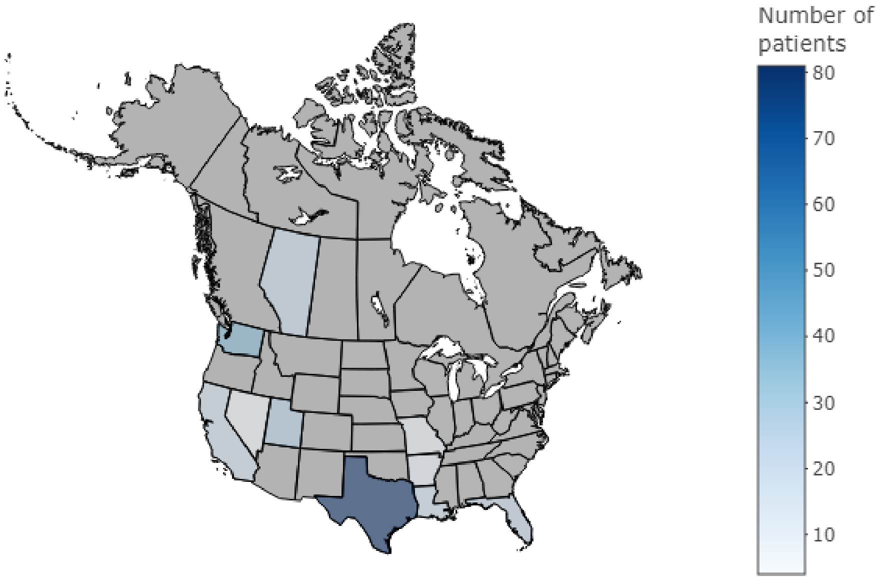

Absolute frequency of patients per state of residence in the USA and Canada.

**Supplemental Figure 4.** Atelectasis percentage on chest CT by obesity category.

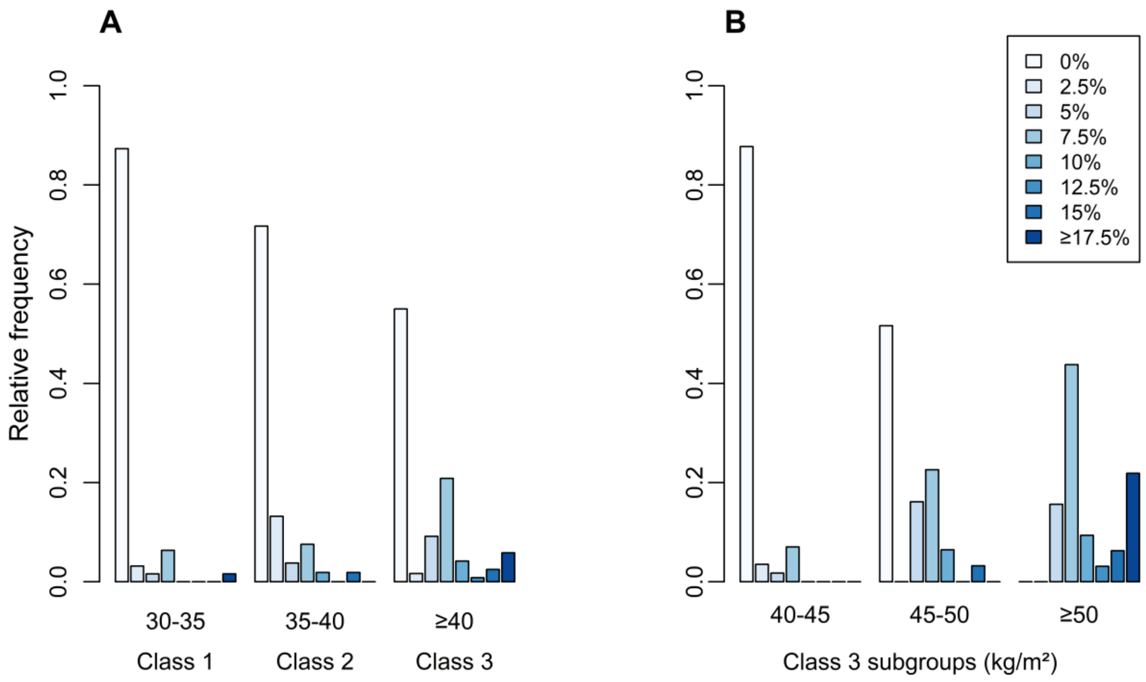

Relative frequency of atelectasis percentage coverage by BMI categories. **A)** World Health Organization obesity class categories. **B)** Class 3 obesity subgroups.

**Supplemental Figure 5.** Preoperative SpO2 predictions for every combination of atelectasis percentage coverage on chest CT and BMI.

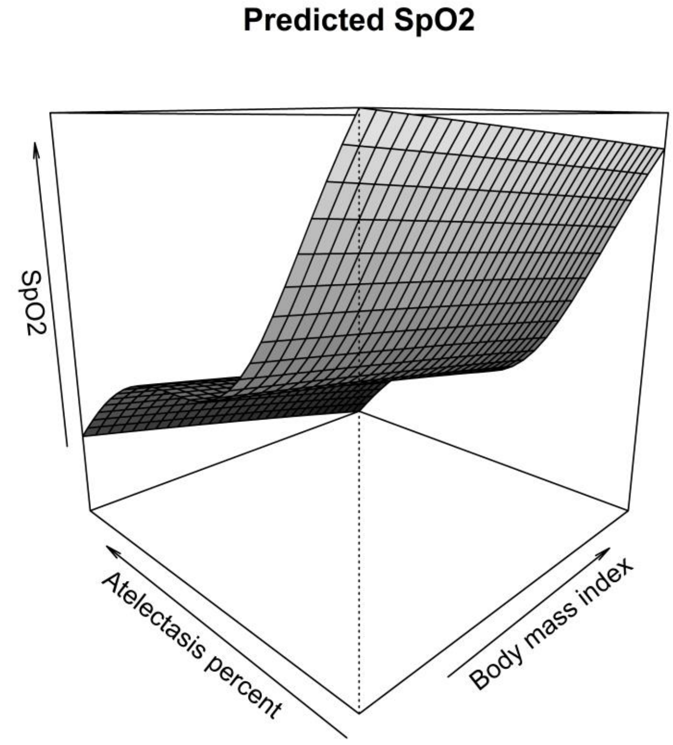

The grid on a gray scale corresponds to predicted values of SpO2 for every possible combination of atelectasis percentage and BMI, weighted for the set of variables *w* (see legend in Supplementary Figure 2 for explanation).

**Supplemental Table 1.** Total, direct (not mediated), and indirect (mediated through atelectasis) effects of body mass index (BMI) on mean SpO2 during the preoperative assessment in the subset of patients with an SpO2 value ≤95%.

| **Characteristic** | **OR** | **95%CI** |
|---|---|---|
| *Total effect of BMI*[1] | | |
| BMI | 0.98 | 0.97—0.99 |
| *Direct and indirect effects of BMI*[2] | | |
| BMI | 1 | 0.99—1 |
| Atelectasis percent | 0.96 | 0.96—0.97 |

Estimates obtained with a generalized linear model (logit link function).

[1]Weighted for the set *L1* (see legend in supplemental figure 2 for explanation).

[2]Weighted for sets *L1* and *L2* (see legend in supplemental figure 2 for explanation) by using the product of weights (*w* = *w1***w2*).

Abbreviations: 95% confidence interval (95%CI), odds ratio (OR)
